# Predicting different dimensions of fatigue from speech data: a longitudinal study in shift workers

**DOI:** 10.1101/2024.06.17.24308769

**Authors:** Agnes Norbury, Alexandra Livia Georgescu, Emilia Molimpakis, Stefano Goria, Nicholas Cummins

## Abstract

**Background:** Shift work, or working outside of normal circadian cycles, is associated with both experience of fatigue and poorer long-term physical and mental health. However, current methods for assessing fatigue present several barriers to building understanding of how these risks develop over time within individuals.

**Objective:** Here, we explore the potential of momentary speech activity-based fatigue measurement in a large, multi-lingual cohort of shift workers, using an intensive longitudinal study design (twice-daily measurement over two weeks in *N*=1,197 individuals from six different countries).

**Methods:** Paralinguistic speech features were used to predict different aspects of acute and chronic fatigue at each study time-point, with performance assessed in unseen (held-out) data. Results are reported both across the dataset as a whole, and for user-specific prediction models.

**Findings:** In the cross-sectional analysis, good (close to or exceeding current state-of-the-art) performance was achieved for both current sleep deprivation and self-reported sleepiness levels. The within-user analysis revealed robust increases in performance, yielding the ability to detect more subjective aspects of fatigue such as pervasive physical and mental exhaustion.

**Conclusions:** These findings offer preliminary support for the utility of using brief momentary speech samples as a low-burden, acceptable, and reliable measure of different aspects of fatigue in high-risk populations such as shift workers.

**Clinical Implications:** Developing brief, accessible measures of different dimensions of fatigue is an important step towards building understanding of how risks for poorer health outcomes develop over time within individuals exposed to significant circadian disruption.

**Summary:** *What is already known on this topic:* Previous studies have indicated that speech data may be a promising source of information about fatigue: however to date these have primarily been carried out in small, unrepresentative samples and in non-naturalistic settings, making results hard to generalize.

*What this study adds:* Here, we present evidence from a large international study where participants both provided momentary speech activity data and reported levels of different aspects of fatigue, as they went about their usual working lives. Importantly, we use a modelling framework that explicitly takes into account potential influences of factors such as age, sex, and language on speech features and target fatigue measures, as well as assessing potential biases and reliability of model output.

*How this study might affect research, practice or policy:* If individuals who are at heightened risk of fatigue-related health problems are able to monitor their fatigue levels regularly and in real-time, this may yield the opportunity to intervene prior to transitioning into less tractable states of poor physical and mental health.

## Background

Fatigue is a pervasive symptom common to a broad range of chronic physical and mental health conditions [1]. Shift work, or working outside of normal circadian sleep-wake cycles, is robustly associated with both elevated fatigue and poorer physical and mental health [2–4]. Multiple inter-related causes of these outcomes have been identified: including increased night-time light exposure and associated circadian rhythm disruption, poor sleep, decreased opportunity for social interaction, and changes to diet and exercise [5, 6]. It is currently less well understood how health risks develop over time within individuals – for example, the tipping points at which shorter-term sleep deprivation or circadian disruption may develop into longer-term exhaustion and physical or mental ill health [7, 8]. The patterns of sleep-wake disruption, fatigue, and other downstream effects experienced by shift workers can be best understood as complex dynamic systems [9]. Building understanding of such systems generally requires repeated measurement of relevant factors in the same individuals over time – particularly if the nature of these processes differs across individuals or people from different socioeconomic groups. However, current methods for assessing fatigue present substantial barriers to this [10].

To date, fatigue has primarily been measured using self-report questionnaires (for example, the Chalder Fatigue Scale [11]) and passive physiological recording methods (electroencephalographic and cardiac activity measures, and video capture of facial expressions or behavioural signals such as yawning; e.g., [12–14]). Whilst questionnaire measures benefit from the validity of probing different subjective aspects of fatigue, they usually concern relatively long timescales (often the last two weeks), and represent a relatively high burden for users under repeated administration (even in brief formats, such as ecological momentary assessment, [15]). On the other hand, some forms of lower-burden passive measurement may be inaccessible at scale, as they require expensive equipment. Further, such tools require sensitive addressing of concerns around privacy, and their acceptability may vary according to factors such as use context, ongoing active consent provision, and the perceived usefulness of output information to individual users [16, 17].

One way of bridging this gap is to use a potentially rich, but accessible and low-cost source of information for fatigue measurement: speech data. Speech production is a complex process, the mechanics of which are known to be affected by many physiological factors related to fatigue, including sympathetic nervous system and hypothalamic–pituitary–adrenal (HPA) axis activity, respiration, and muscle tension [18]. Several previous studies have detected signal for fatigue in paralinguistic speech features, i.e. non-verbal communication features that accompany speech, including pitch (e.g., [19–21]. However, these studies have primarily described small samples of individuals in non-naturalistic settings (for example, experimentally-evoked sleep deprivation in otherwise healthy participants) [22].

While there is no universally accepted definition of fatigue [23], it is generally accepted that it is a multi-faceted construct, encompassing physical, cognitive, and emotional impacts across multiple time-scales [24]. For example, some questionnaire measures of fatigue distinguish between acute tiredness and symptoms of prolonged or chronic fatigue [25]. Whilst the former are short-lived and recoverable with rest, the latter include experience of negative emotions, concentration problems, and motivational deficits which impact everyday functioning. Accounts from people living with significant fatigue (including shift workers) stress that their experience includes not only disturbed sleep, but effects such as a sense of ingrained tiredness, pervasive lack of energy, and broader challenges to physical and psychological well-being [26–28]. Both within shift workers, and more generally, these broader aspects of fatigue have been found to correlate poorly with self-reported sleepiness, suggesting they reflect at least partially separable underlying processes [22, 29].

In summary, while speech analysis has shown promise for detecting physiological states related to fatigue, existing studies do not provide strong evidence for the effectiveness of this approach for both more acute aspects of fatigue (such as current sleep deprivation state and sleepiness levels) and broader subjective impacts (such as pervasive physical and mental exhaustion, general lack of energy, and motivational difficulties). Further, to date, research effort in this area faces substantial barriers to real-world implementation: given reliance on small or unrepresentative samples. The ability to accurately measure different aspects of fatigue, using a low-burden, acceptable, affordable and reliable measurement tool would therefore have strong translational impact. In particular, if individuals who are at heightened risk of fatigue-related health problems are able to monitor their fatigue levels regularly and in real-time, this would yield the opportunity to identify meaningful trends over time, and potentially intervene prior to transitioning into less tractable states of poor health [8].

## Objectives

Here, we explore the potential of speech-based fatigue measurement in a large, multi-lingual cohort, with data collected under an ecologically valid setting (as participants went about their usual working lives). We tackle the issue of the multidimensionality of fatigue by explicitly examining to what extent different aspects of fatigue (from acute sleep deprivation state to chronic motivational difficulties) may be detectable in speech data. Importantly, we acknowledge that speech is a variable signal that is known to be affected by factors like sex, age, and accent. Failure to account for this when predicting target variables that may also be systematically related to these factors may lead to over-optimistic estimates of speech model performance [30, 31]. Since we have reason to believe that aspects of fatigue such as mental exhaustion or loss of motivation may be non-trivially associated with participant demographics (e.g., [32]), we use a modelling approach designed to appropriately partition variance due to demographic differences across individuals, and report model performance separately for different groups. Finally, we explore the acceptability of speech-based activities for repeated longitudinal tracking of fatigue, within the same individuals.

## Methods

### Ethical approval

Study procedures were reviewed by an independent research ethics expert (Dr. David Carpenter) working under the auspices of the Association of Research Managers and Administrators (ARMA, https://arma.ac.uk/) and approved on 22nd November 2023. All participants gave written informed consent.

### Recruitment

Participants were recruited using an online research participation platform (Prolific, https://www.prolific.co/), and were required to be 18-65 years old, free from neurological or language disorders and vision and hearing problems, and to speak English or Spanish as their first language. Participants were also required to be currently employed in a shift-work pattern. In the later stages of the study, a small number of recruitment waves targeting regular 9-5 workers were also included, in order to provide a comparison group and ensure a wide range of observed work-related fatigue patterns.

### Study protocol

The study was carried out according to an intensive within-user longitudinal design (Figure 1A). All activities were carried out remotely on participants’ own devices, using the thymia research platform [33]. Briefly, participants were prompted to complete study activities twice a day for a two-week period, once before the start and once after the end of their work shift.

**Figure 1:**
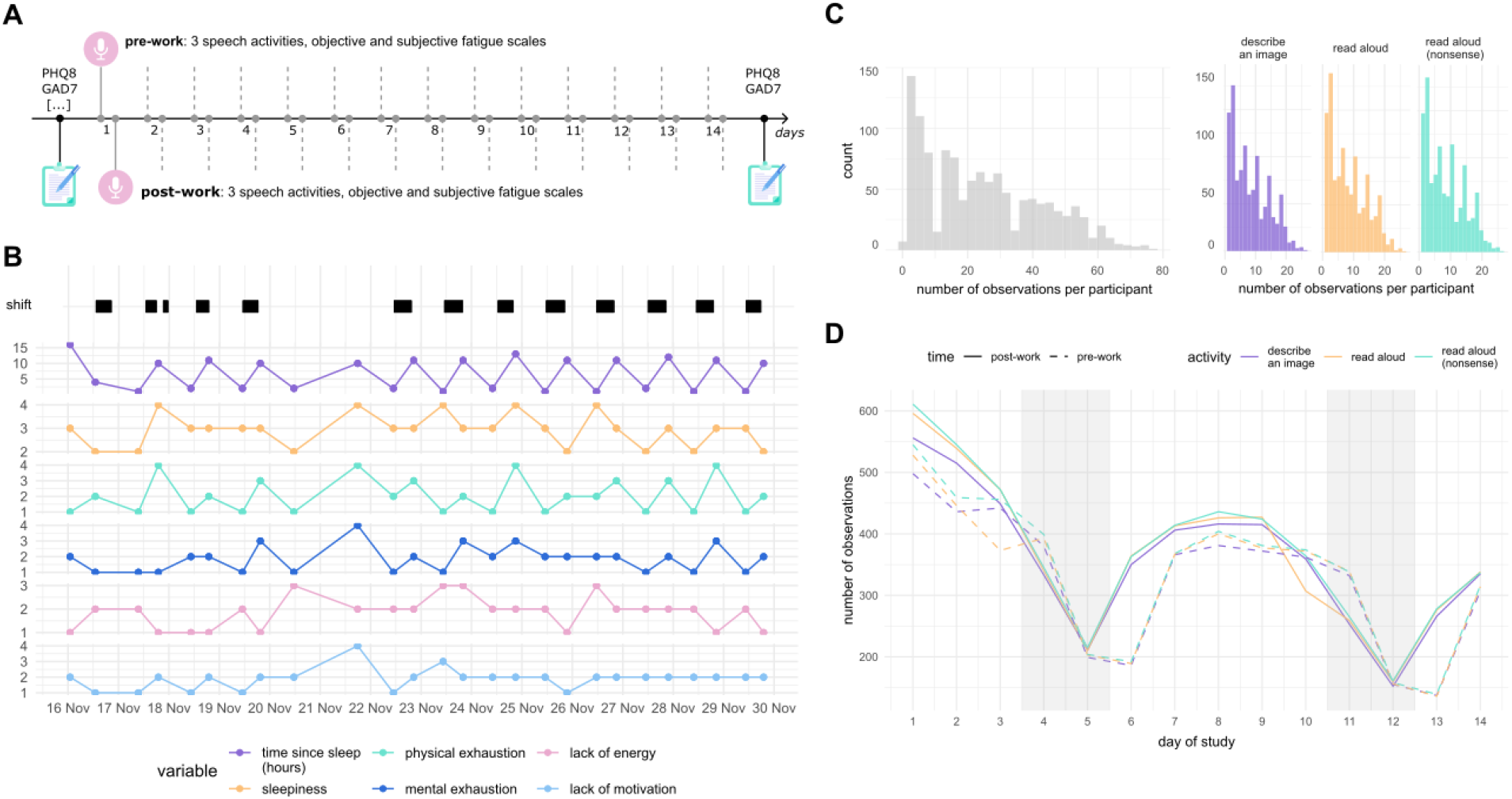
Overview of study design and available speech data. **A** Study participants were prompted to complete study activities twice a day for a two-week period, once before and once after completing their work shift. At each time-point, participants were asked to complete three brief speech activities and provide information about their work, sleep, and fatigue levels. Physical and mental health symptoms (including the Patient Health Questionnaire, PHQ-8, and the Generalised Anxiety Disorder questionnaire, GAD-7) were measured at the start and end of the study. **B** Work, sleep, and fatigue data from an example participant, over the two-week study period. Top row, reported shift onset and offset times (with black bars representing time at work); second row, sleep deprivation status (hours since last sleep); further rows, self-reported fatigue levels. **C** Distributions of available number of speech recordings following preprocessing and data cleaning per participant (left), and by participant and speech activity (right). **D** Total number of speech recordings available by study day and time-point (pre *vs* post-work) and speech activity type. Grey shading represents weekends.

At each study time-point, participants were requested to complete a set of activities, of approximately 10 minutes total length. Activity completion reminders were sent out twice a day during the study period, with each session available for 12 hours to cover the full 24-hour period. If participants were not able to complete activities within this 12-hour window, they were instructed to wait until the next available session and corresponding shift. Requesting that participants complete study activities relative to their own unique work patterns, rather than at set times of day, allowed us to maximise variation in observed work-related fatigue, independently of circadian time.

At the start and end of the study, participants provided detailed demographic and relevant clinical information – including their recent experience of physical and mental health symptoms (see below and Supplementary Methods).

### Measures

Here, we analyse data from three different commonly-used speech elicitation activities (varied text-reading and image description tasks, all ~ 1 minute in length; see Supplementary Methods), and self-reported work, sleep, and subjective fatigue levels.

At each time point, participants reported the time their last shift ended and the time when they most recently woke from sleep. They also recorded their current fatigue levels across several different dimensions using 10-point likert scales: specifically, sleepiness, physical exhaustion, mental exhaustion, lack of energy (anergia) and lack of motivation (for full item descriptions see Supplementary Methods). These dimensions were chosen based on a review of the previous fatigue literature, including existing self-report measures and lived experience accounts (see Background). Fluctuations in work, sleep, and fatigue for an example participant over the course of the study are plotted in Figure 1B.

Participants’ experience of symptoms of depression and anxiety over the last two weeks was assessed at the start and end of the study using brief measures suitable for use in the general population (the PHQ-8 and GAD-7; [34, 35]).

### Speech data preprocessing and data cleaning

Audio recordings from each activity type were preprocessed according to the following steps. First, recordings were run through a Speech To Text engine (AWS Transcribe) to produce a JSON file with word by word transcription and time-stamps. Using the time-stamps from the transcription file, trailing silences were removed. Utterance cuts (snippets of speech containing pauses shorter than 0.5 seconds) were then created from the audio data, and the different utterance cuts joined together with a silence of 0.2 seconds. Finally, from the joined utterance cuts, the first 20 seconds of audio data (16Khz sampling rate) were selected for further analysis.

If the audio quality did not allow for transcription, or the final audio snippet was shorter than 20 seconds, the recording was excluded from further analysis. In order to focus our analysis on samples collected according to the intended study protocol, recordings for which time since last sleep or time since last shift was reported to be greater than 24 hours were also excluded.

### Speech feature generation

In order to transform audio snippets into a feasible format for simple supervised learning models, we generated embeddings using pre-trained large models specialised for paralinguistic tasks. Specifically, the 20 second audio snippets were fed to a TRILLsson model [36], which produces an embeddings vector of length 1024 for each snippet. The TRILLsson model family has previously shown strong performance in paralinguistic tasks spanning from emotion recognition to Alzheimer’s detection [37]. Within the TRILLsson family we selected the model with the highest reported performance (TRILLsson5).

### Statistical analysis

In order to confirm that our dataset contained participants with significantly disrupted sleep-work patterns and elevations in other fatigue measures, we first compared sleep/work status and self-reported fatigue levels at each time-point between shift and non-shift workers.

We then sought to determine whether features derived from the brief speech activities carried signal for fatigue across the dataset as a whole. Following previous observations that more objective measures of fatigue (e.g., sleep deprivation state, sleep onset latency) may be differently recoverable from speech data than more subjective measures (e.g., self-reported sleepiness) [38], we report results separately for time since sleep/work and other fatigue measures. Given the longitudinal nature of the data, we next examined potential gains in performance by including past observations from the same user in prediction models.

Descriptive and initial statistical analyses were carried out in R 4.3.2 (R Foundation for Statistical Computing, 2023). Prediction analyses were carried out using python 3.12.2 (Python Core Team, 2024).

#### Analysis of demographic and clinical data

Differences between shift and non-shift workers in within-study fatigue and baseline physical and mental health were assessed using (generalized) linear models as implemented in the R package lme4. Although our sample predominantly consisted of shift workers, these models are relatively robust to heterogeneity in sample size and variance between groups. Where multiple observations were available per participant (i.e., fatigue measures), data were analysed using mixed-effect models with a random intercept per participant. All models included participant age and gender identity as covariates. Reported effect sizes are *F* statistics (df numerator, df denominator), which quantify the relative fit of a model including a given independent predictor, compared to one without it.

#### Predicting fatigue from speech data

Prior to further analysis, the preprocessed speech dataset was split into training and test splits in a 0.9:0.1 ratio. Splits were stratified by user ID (no shared users across splits), and approximately balanced for birth sex, language, and study wave (country of recruitment and shift work status).

#### General methods

Fatigue measures were predicted from the set of TRILLsson5 voice features using L2-regularized regression models. Although more complex models are often employed in speech feature analysis, regularized linear models often perform well in high-dimensional prediction problems, and carry the advantage of being easily interpretable, compared to some non-linear models [39]. [if we go with the averaged model, need to describe the averaging here]. For each model, degree of regularization was determined using nested 5-fold cross-validation.

All models were fit to training data using a mixed-effects framework, as implemented in the python package MERF. Mixed-effect machine learning methods have previously been shown to provide improved predictive performance compared to standard approaches in cases where random-effects are non-neglible [40, 41] – as we might expect to be the case for some subjective fatigue measures (see Background). Briefly, this approach aims to dissociate between fixed and random effects on target variables using iterative calls to the relevant model (here, ridge regression) within the context of a supervising expectation-maximization (EM) algorithm. At each iteration, a standard prediction model is built from transformed data, where the current estimate of the random-effects component has been removed. The covariance of the distributions from which random-effects weights are drawn is learned over iterations, with convergence assessed using the generalized log-likelihood criterion [40]. If a new observation belongs to a previously seen cluster (here, user ID), both fixed and random-effects estimates are used for prediction. If a new observations belongs to a previously unseen cluster, only the population-averaged fixed effect estimates are used.

Age category (18-32, 32-65), birth sex (male, female), language (English-GB, English-US, English-Other, and Spanish) and speech activity type (read aloud, read aloud: nonsense, and describe an image) were used as random effects in all models, given evidence that these factors may systematically affect voice features [2].

In order to assess the impact of amount of available speech data at each time-point, we report performance for both the model described above (where individual speech activity feature vectors were treated as independent predictors of user fatigue at each time-point), and a model where speech features were first averaged across activities for each user at each time-point (yielding a single speech feature vector for each user at each time-point).

Model performance was assessed using Spearman’s *ρ* correlations between predicted and observed values in held-out or independent test set data, alongside the root mean squared error (RMSE) of predictions. As a benchmark, we considered performance of the winning model from the 2019 Interspeech competition for predicting self-reported sleepiness from the Düsseldorf Sleepy Language Corpus [42] (*ρ*=0.387) and several subsequent replication attempts (Pearson’s *r*s=0.325, 0.367, 0.365, 0.383; see [38]). We therefore assumed *ρ*>0.36 (the average of these values) to be indicative of good performance.

#### Cross-sectional analysis

Time since sleep, time since work, and other self-reported fatigue measures were first predicted cross-sectionally across all study time-points using a mixed-effects framework, with observations clustered by user ID and random effects as described above. Since test data contained only unseen participants, predictions were made using the population-averaged fixed effect estimates. For cross-sectional analyses, we report mean (SD) performance across nested cross-validation folds in the training set, followed by performance in the independent test set.

#### Reliability analysis

In order to assess the reliability of model predictions across speech activities, intra-class correlation coefficients (ICCs) were calculated for concordance between predictions from two different speech activities at each time-point, using the ICC function from the R package psych. We report both ICC(3,1) and ICC (3,k) [43] to provide a measure of both the single speech activity reliability and average reliability across measures (given relevance for likely use-cases where users complete *>*1 speech activity at each assessment).

#### Assessment of bias

We report predictive accuracy broken down by age, sex, and language/accent categories, in order to identify any differences in performance across these groups and potential limits to generalizability of findings.

#### Within-user analysis

In order to assess the potential benefits of constructing within-user prediction models, datasets were filtered for participants with speech samples from more than 6 different study time-points (*N*=636 participants; for a comparison between users included and excluded from this analysis see Supplementary Results). Models were trained on data from a randomly selected subset of time-points from each user, with performance assessed on an unseen time-point from the same user. Specifically, we assessed performance for inclusion of data from 1-7 different time-points per user in group-level mixed-effect models for each target. Random selection of the train:test time-point splits was repeated 10 times for each model, and mean (SD) performance across iterations reported. Since these models included participant-level random-effects (the same cluster IDs were present in train and test splits), we compared performance of the speech data models to null models which only had access to user cluster IDs and random-effect covariate values (in essence, an intercept-only model).

### Findings

#### Participants

Study participants (*N*=1,197) are described in Table 1. The majority of participants (86%) reported English as their first language, for the remaining participants (14%) their first language was Spanish. Where information was available (81% of participants), 99% reported that their first language was their daily language. 83% of participants were shift workers, of which 43% worked a shift pattern that included night shifts. The most common shift work field was healthcare and social assistance.

**Table 1:** Description of study participants. Unless otherwise specified, values represent N (%) of valid data. Data were complete for all demographic, language, and shift work status variables, with a small proportion of missingness for other work pattern variables (mean=7%). *PHQ-8*, Patient Health Questionnaire 8-item measure of depression symptoms; *GAD-7*, Generalized Anxiety Disorder questionnaire.

| Variable |  | Value |
| --- | --- | --- |
| Age (years) | mean (SD) | 33.6 (9.9) |
|  | range | 18-65 |
| Gender | Women | 642 (53.6%) |
|  | Man | 533 (44.5%) |
|  | Non-binary | 22 (1.8%) |
| Birth sex | Female | 662 (55.3%) |
|  | Male | 534 (44.6%) |
|  | Prefer not to say | 1 (0.1%) |
| Race/ethnicity | Asian | 26 (2.2%) |
|  | Black | 39 (3.3%) |
|  | Mixed | 130 (10.9%) |
|  | Other | 59 (4.9%) |
|  | White | 943 (78.8%) |
| Country of residence | Australia | 6 (0.5%) |
|  | Canada | 31 (2.6%) |
|  | Ireland | 9 (0.8%) |
|  | Mexico | 157 (13.1%) |
|  | Spain | 61 (5.1%) |
|  | United Kingdom | 711 (59.4%) |
|  | United States | 222 (18.5%) |
| First language | English | 1027 (81.3%) |
|  | Spanish | 170 (14.2%) |
| Shift worker | Yes | 988 (82.5%) |
|  | No | 209 (17.5%) |
| Work pattern | Regular 9-5 | 201 (23.8%) |
|  | Rotating shifts not including night shifts | 258 (30.5%) |
|  | Rotating shifts including night shifts | 342 (40.5%) |
|  | Permanent night shifts | 44 (5.2%) |
| Work hours per week | Less than 35 | 236 (21.2%) |
|  | 35-39 | 394 (35.4%) |
|  | 40-49 | 411 (37.0%) |
|  | 50+ | 71 (6.4%) |
| Shift length<br>(shift workers only) | 8 hours | 397 (49.1%) |
|  | 10 hours | 141 (17.5%) |
|  | 12 hours | 172 (21.3%) |
|  | 14 hours | 13 (1.6%) |
|  | 14 hours+ | 11 (1.4%) |
|  | Other | 74 (9.2%) |
| Shift work industry<br>(shift workers only) | Healthcare and social assistance | 230 (28.6%) |
|  | Manufacturing | 31 (3.9%) |
|  | Military | 11 (1.4%) |
|  | Services | 146 (18.2%) |
|  | Transportation | 47 (5.8%) |
|  | Warehousing | 13 (1.6%) |
|  | Wholesale and retail trade | 121 (15.0%) |
|  | Other | 213 (26.4%) |
| Health suffers due to work | No | 669 (60.1%) |
|  | Yes | 445 (39.9%) |
| Chronic physical illness | No | 891 (77.4%) |
|  | Yes | 255 (22.2%) |
|  | Prefer not to say | 5 (0.4%) |
| PHQ-8 total | mean (SD) | 8.4 (5.2) |
| GAD-7 total | mean (SD) | 6.8 (5.0) |

#### Available speech data

The cleaned and preprocessed speech dataset consisted of 29,525 individual voice recordings. The mean number of voice recordings per participant (over all three voice activities per time-point) was 24.7 (SD 18.0). For distributions of available speech samples by user and study time-point see Figure 1C,D.

The training set consisted of 26,679 observations from *N*=1,076 users, and the test set 2,846 observations from *N*=121 users (no users overlapped between data splits).

#### Shift work status, fatigue, and physical and mental health

##### Association between shift work status and fatigue

Over the course of the study, shift workers reported disrupted sleep and rest from work patterns compared to non-shift workers (higher mean time since sleep, lower mean time since shift, *F*_1,666_ = 14.1*, F*_1,743_ = 35.7, both *p <* 0.001; Figure S1A).

Shift workers also showed substantially elevated subjective fatigue levels, across all self-report scales (sleepiness, physical exhaustion, mental exhaustion, lack of energy and lack of motivation: *F*_1,970_ = 83.1*, F*_1,955_ = 94.4*, F*_1,976_ = 49.4*, F*_1,988_ = 43.9*, F*_1,1032_ = 26.9; all *p <* 0.001; Figure S1B). These effects persisted in models controlling for time since sleep and time since shift at each time-point, suggesting the presence of distinct cumulative (rather than situation-responsive) subjective fatigue patterns in this group (*F*_1,974_ = 71.6*, F*_1,968_ = 80.3*, F*_1,986_ = 38.0*, F*_1,773_ = 37.1*, F*_1,1034_ = 24.5; all *p <* 0.001).

##### Association between shift work status and physical and mental health

Shift workers were more likely to report that their health suffered due to their work (*z*_1107_ = 3.86*, p <* 0.001*, OR* = 1.97), and that they suffered from a chronic physical illness (*z*_1139_ = 2.02*, p* = 0.04*, OR* = 1.51).

Shift workers had higher baseline PHQ-8 total depression scores (*F*_1,1190_ = 22.3*, p <* 0.001), and showed a non-significant trend towards higher GAD-7 total anxiety scores (*F*_1,1190_ = 2.9*, p* = 0.088) (Figure S1C). Across self-reported depression symptoms, the greatest elevations were evident for PHQ-8 items 4 (“*Feeling tired or having little energy*”), 5, (“*Poor appetite or overeating*”), 3 (“*Trouble falling or staying asleep, or sleeping too much*”), and 7 (“*Trouble concentrating on things*”; mean differences = 0.44, 0.35, 0.31, 0.28; Figure S1D).

Within shift workers, a work pattern that included night shifts and working in healthcare and social assistance, but not long hours in general (40+ hours per week), were associated with higher total depression scores (*F*_1,505_ = 4.56, 12.8, 0.180; *p* = 0.033*, p <* 0.001*, p* = 0.672).

Our cohort therefore consisted primarily of shift workers, who reported significantly elevated fatigue and poorer physical and mental health compared to regular 9-5 workers. Next, we sought to examine whether these features were detectable in momentary speech activity data.

#### Predicting fatigue from speech data

##### Cross-sectional analysis

###### Time since sleep and time since work

Across users and time-points, good performance was achieved for predicting time since sleep from speech data (for the single activity model, nested cross-validated training set performance: mean *ρ* = 0.360 (SD 0.018); test set performance: *ρ* = 0.338; for the averaged activity model, cross-validated performance: mean *ρ* = 0.371 (SD 0.025); test set performance: *ρ* = 0.365) (Table 2). Time since last shift was harder to detect using the same model of speech features (single activity model train, test: *ρ* = 0.254 (0.015), *ρ* = 0.203; averaged activity train, test:*ρ* = 0.259 (0.026), *ρ* = 0.212). By comparison, a simple linear regression model containing only demographic factors included as random-effects in the speech model as predictors performed poorly in unseen data (time since sleep: *ρ* = 0.065, 0.002; time since shift: *ρ* = − 0.007, 0.041). Histograms showing the distribution of observed time since sleep and time since shift values in the training data are available in Figure S2A.

**Table 2:** Summary of cross-sectional model performance. Train, performance in held-out data (mean, SD across cross-validation folds. Test, performance in independent test set. *ρ*, Spearman’s correlation coefficient between predicted and observed values. *RMSE*, root mean square error of predictions (for time variables, in hours [possible range 0-24], for self-report variables in likert scale units [possible range 0-10]).

|  | single speech activity models |  |  |  | averaged over speech activities models |  |  |  |
| --- | --- | --- | --- | --- | --- | --- | --- | --- |
|  | train |  | test |  | train |  | test |  |
| | $\rho$ | *RMSE* | $\rho$ | *RMSE* | $\rho$ | *RMSE* | $\rho$ | *RMSE* |
| time since sleep | 0.360 (0.018) | 5.84 (0.052) | 0.338 | 5.96 | 0.371 (0.025) | 5.74 (0.180) | 0.365 | 5.78 |
| time since shift | 0.254 (0.015) | 7.02 (0.171) | 0.203 | 7.56 | 0.259 (0.026) | 6.97 (0.122) | 0.212 | 7.44 |
| sleepiness | 0.334 (0.020) | 2.13 (0.060) | 0.254 | 2.09 | 0.345 (0.037) | 2.12 (0.049) | 0.265 | 2.08 |
| physical exhaustion | 0.256 (0.050) | 2.52 (0.047) | 0.138 | 2.59 | 0.269 (0.048) | 2.51 (0.041) | 0.158 | 2.59 |
| mental exhaustion | 0.234 (0.054) | 2.52 (0.046) | 0.133 | 2.49 | 0.258 (0.056) | 2.50 (0.040) | 0.172 | 2.47 |
| lack of energy | 0.257 (0.058) | 2.50 (0.088) | 0.153 | 2.50 | 0.272 (0.049) | 2.49 (0.027) | 0.185 | 2.49 |
| lack of motivation | 0.190 (0.065) | 2.51 (0.102) | 0.146 | 2.32 | 0.225 (0.056) | 2.50 (0.052) | 0.171 | 2.33 |

###### Subjective fatigue measures

Cross-sectionally, the strongest prediction results were observed for current sleepiness (single activity model train, test: *ρ* = 0.334(*SD* = 0.020), *ρ* = 0.254; averaged activity model train, test: *ρ* = 0.345 (SD=0.037), *ρ* = 0.265) (Table 2). Of note, as well as being theoretically distinct from sleep deprivation state (see section), time since sleep and subjective sleepiness ratings were only moderately correlated across observations (*ρ* = 0.258*, p <* 0.001, paired-samples rank correlation test).

Other subjective fatigue measures were harder to detect across speech samples (for single activity models, physical exhaustion: *ρ* = 0.256(0.05), 0.138; mental exhaustion: *ρ* = 0.234(0.05), 0.133; lack of energy: *ρ* = 0.257(0.06), 0.133; lack of motivation *ρ* = 0.190(0.07), 0.146; for averaged activity models, physical exhaustion: *ρ* = 0.269(0.05), 0.158; mental exhaustion: *ρ* = 0.258(0.06), 0.172; lack of energy: *ρ* = 0.272(0.05), 0.185; lack of motivation *ρ* = 0.225(0.06), 0.171). As expected, subjective fatigue measures were more strongly predicted by a demographics-only linear regression model than time since sleep and work measures (sleepiness: *ρ* = 0.126, 0.164; physical exhaustion: *ρ* = 0.109, 0.144; mental exhaustion: *ρ* = 0.060, 0.184; lack of energy: *ρ* = 0.101, 0.168; lack of motivation: *ρ* = 0.109, 0.160). Histograms showing the distribution of observed time since sleep and time since shift values in the training data are available in Figure S2C.

###### Reliability

Concordances between predictions based on different speech activity data at each time-point were high, suggesting that performance relies on features of the model pipeline not specific to a given activity type. Specifically, for time since sleep, ICC(3,1) (reliability of a single activity measurement) was 0.804 [95% CI 0.779,0.826], and ICC(3,k) (reliability of the average across activity measurements) was 0.891 [0.876,0.905](read aloud *vs* describe an image activity predictions). For time since shift, ICC(3,1) was 0.814 [0.790,0.835] and ICC(3,k) was 0.897 [0.883,0.910]. For subjective fatigue ratings, ICCs were sleepiness: 0.853 [0.834,0.870], 0.921 [0.909,930]; physical exhaustion: 0.859 [0.841,0.875], 0.924 [0.913,0.933]; mental exhaustion: 0.836 [0.814,0.854], 0.910 [0.898,0.921]; lack of energy: 0.872 [0.856,0.877], 0.932 [0.922,0.940]; lack of motivation: 0.856 [0.837,0.872], 0.922 [0.911,0.932].

###### Bias assessment

For time since sleep and sleepiness fatigue measures, cross-sectional model performance estimates were relatively robust across sex and language/accent groups (see Table S1, Table S2). In general, performance was better for groups with higher representation in the training dataset (<32 years of age, and female sex). For the subjective fatigue measures physical exhaustion, mental exhaustion, lack of energy and lack of motivation, performance was poor for individuals from the en-OTHER language/accent category (i.e., Irish, Australian, and Canadian English speakers) - likely due to the low representation of this group in the training data (227 observations from 37 users).

##### Within-user analysis

We next tested whether performance could be improved by including previous observations from the same user in the training data set, and how many observations this requires.

###### Time since sleep and time since work

Robust improvements in predictive accuracy were observed after inclusion of several previous training time-points per user. At inclusion of three training time-points, accuracy for time since sleep increased to 0.402 (SD across random train-test splits 0.017) for the single activity model and 0.408 (SD 0.028) for the averaged across activity model. Asymptotic performance (at inclusion of seven time-point) was 0.507 (0.31) for the single activity model and 0.525 (0.031) for the averaged activity model (Figure 2; Table S3, Table S4). For time since shift, performance after four time-points was 0.314 (0.025) and 0.304 (0.020) for the single activity and averaged activity models, and after seven time-points reached 0.355 (0.024) and 0.370 (0.038), respectively.

**Figure 2:**
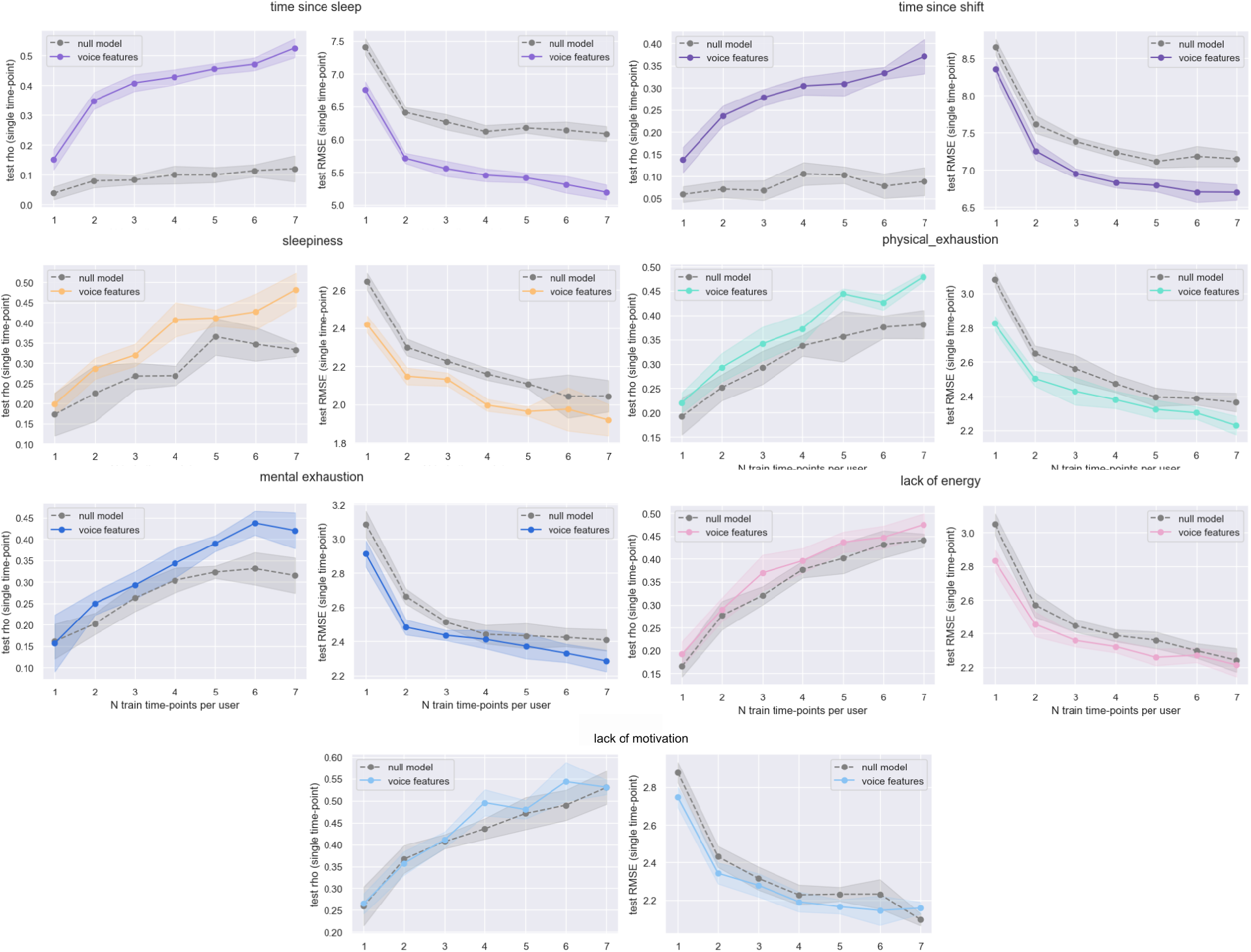
Within-user model performance. For each fatigue measure, performance for a single unseen time-point is plotted against number of observations from the same user included in the training data. Speech feature models included all speech features at each time point, cluster (user) ID, and random-effects. Null models included only cluster (user) ID and random-effects values. Rho values are Spearman correlation coefficents between predicted and observed values in independent test set data. RMSE, root mean squared error of predictions. Results plotted here are for the averaged across speech activity model, for results from the single activity model please see Figure S4.

Please note, the single time-point accuracy here is lower than that reported for the cross-sectional analysis due to the decrease in number of training observations (and users) that resulted from filtering the dataset for a minimum number of speech data time-points. Under a mixed-effects framework, a higher number of unique clusters (here, users) has previously been found to be associated with increased predictive accuracy [44].

###### Subjective fatigue measures

For the subjective fatigue measures, assessing the contribution of speech data to predictive accuracy was more challenging, due to strong between-participant effects in mean ratings, and lower variance in observed values over the study time-scale (see Supplementary Results; Figure S3A). These features of the data meant that a null model containing only user identify and random effects (in essence, predicting values by fitting an intercept to previous observations for each user) performed very well for some scales (asymptotic *ρ* for lack of motivation ∼0.70, for sleepiness, ∼0.50; Figure S3B).

In order to be able to assess potential contributions of speech features to within-user prediction models for these measures, we therefore further filtered the data for a minimum SD across subjective fatigue ratings in included time-points of 1.0 (new *N* users=488). In this subset of the data, the null (random-effects only) model still performed relatively well, but with lower performance ceilings (asymptotic *ρ* for lack of motivation ~0.50, for sleepiness, ~0.35; Figure S3C).

In this subset, a model including speech features was able to out-perform the null model after 2-3 training time-points for sleepiness (after 3 time-points, single activity model *ρ*=0.325 (0.049), averaged activity model *ρ*= 0.320 (0.027); with asymptotic performance approaching 0.45, 0.48, respectively) (Figure 2; Table S3, Table S4). For physical and mental exhaustion, performance of the speech feature model was superior to the null model after 3-4 observations (after 4 observations, single activity model *ρ*s=0.392 (0.029), 0.367(0.024), averaged activity model *ρ*s= 0.373 (0.030), 0.343 (0.035)). For the most slowly moving subjective ratings, lack of energy and lack of motivation, the speech model did not convincingly out-perform the random-effects only models - likely at least partly due to fundamental limitations in the nature of this data over the study time-period.

##### Speech activity acceptability

Given a key motivation for this work is to develop methods for longitudinal tracking of fatigue within individuals, we also explored measures of speech activity acceptability in study participants. This is an imperfect way of assessing the potential usability of a future speech-based fatigue-tracking tool, given the research study setting (more intensive and demanding measurement schedule than may be expected in translational settings), and that study feedback measures only available in the subset of users who completed the study. In the data available (*N*=738 out of 1,197 users completed the final assessment), overall study liking ratings were generally high (mean rating=5.98 [SD 1.1], on a scale from 1 [*did not like the study at all*] to 7 [*liked it extremely*]). Most users who completed the study also reported a strong preference for continuing to use the platform for another two weeks, if it was available (mean rating=6.17 [SD 1.3], on a scale from 1 [*not at all likely*] to 7 [*extremely likely*]).

## Discussion

Here, we explored the potential of using momentary speech data samples to track different aspects of fatigue in a cohort of individuals with predominant sleep/work cycle disruption (shift workers). When considering the dataset as a whole, we found evidence of good (close to or exceeding current state-of-the-art) performance for predicting both time since sleep and current sleepiness levels, using relatively simple and interpretable linear models of paralinguistic speech features. This is notable, as not only were these two fatigue measures only weakly correlated in our large multi-lingual dataset, but they are considered to reflect different underlying processes. Specifically, sleepiness is thought to reflect not just current sleep deprivation levels, but an alteration or imbalance in competing sleep/wake mechanisms [23]. We found slightly superior performance for models that averaged across all available speech activities for each user at each time-point (possibly, increasing signal:noise ratio), with performance of a single speech activity models being only ∼10% lower.

Other fatigue measures (physical and mental exhaustion, lack of energy, and lack of motivation) were both more strongly biased by individual demographics, and harder to accurately detect in a cross-sectional analysis. It is possible that lower performance for these measures was the result of our choice of a modelling framework that appropriately accounts for variance in target variables systematically related to covariates that may be recoverable from speech data (i.e., lower true signal in speech features after removing variance in target fatigue measures related to factors such as age and sex) [30, 31]. It is also possible that features of the data recorded for these measures at our chosen study time-scale made them harder to predict. Specifically, whilst study design was optimized to induce maximal variation in time since sleep and work variables, we observed significantly lower variance in measures quantifying subjective experiences of fatigue. In the future, longer time-scales (>2 weeks) may be required to collect training data with greater variance for these more slow-moving or chronic aspects of fatigue.

A key question in machine learning analysis of speech features is what the models are actually learning. For example, a recent analysis of the Düsseldorf Sleepy Language Corpus concluded that features of the data make it hard to learn features of sleepiness without at the same time learning features of speaker identity [45]. We attempted to limit such effects in our analysis by both using a study design that attempted to maximise within-user variance in current fatigue levels and by using a mixed-effects framework that explicitly partitions within vs between-user variance in observed data. We also explored whether models trained on different numbers of speech activity observations would show improved performance for predicting unseen fatigue measures from the same set of users. Using this approach, we found that after inclusion of 5 previous observations per user, performance reached good levels for several different aspects of fatigue (for the averaged across activity models: time since sleep, *ρ*=0.45; sleepiness, *ρ*=0.41; physical exhaustion, *ρ*=0.44; mental exhaustion, *ρ*=0.39). Of note, for all these measures, a model including speech features out-performed the null (intercept-only) model, suggesting contribution of speech data beyond more trivial prediction on the basis of previous target values for each user. This suggests there may be substantial utility in including previous observations from individual users and a modelling framework that includes user-identity information for predicting more subjective or chronic dimensions of fatigue.

Generation of reliable predictions is a prequisite for both accurate detection of individual differences at any give time, and ability to measure changes in values over time in the same users. However, the test-retest reliability of model output is often unreported for commercial AI-based digital assessment tools [46]. This is important, given the concordance of some speech features across speech activity types and time has previously been reported to be low (median values between 0.02 and 0.55; [47]). Here, we found that predicted fatigue scores were highly reliable between different speech activities at the same time-point (median ICC for single measurement = 0.804, for average across measurements = 0.891). ICC values of 0.75 and above are usually recommended for clinical-grade applications [48].

We also assessed the potential for bias in model outputs by separately reporting performance accuracy according to different demographic splits of the data. Whilst cross-sectional performance was acceptable for time since sleep and sleepiness fatigue measures was relatively strong across age, sex, and language/accent categories, there was some evidence of biasing of performance towards individuals with age groups and birth sexes more highly represented in the training data (i.e., younger females).

Finally, we attempted to gauge the level of acceptability of brief speech activities for measuring fatigue in our study participants. Previous research has found users to be generally open to the idea of using speech recordings to monitor sensitive states such as mental welfare, particularly if concerns around privacy are adequately addressed [17]. Here, our analysis is somewhat limited in scope by both differences in likely implementation between research study and future translational settings (higher burden, but compensated participation) and selection bias (only participants who reached the study end-point completed post-study acceptability ratings). 738/1,197 or 62% of participants included here completed the post-study assessment. In these participants, overall study liking and willingness to continue using the platform ratings were generally high. In free-text responses, several participants described the speech activities as engaging, which may have encouraged higher continued engagement than some intensive self-report protocols (which often report steep drop-offs in participation rates over studies of less than two weeks duration [15, 49]). Ensuring minimal participation burden may be particularly important for maintaining engagement in populations experiencing significant levels of fatigue and motivational difficulties. Future work should explore user experience in greater detail, in order to support user-centred design of speech-based fatigue monitoring tools. For example, it has previously been shown that people with depression are less comfortable with free-speech than scripted reading tasks [50].

Our study has some general limitations. Although a strength of the dataset was that it describes a large, international and multilingual sample, study exclusion criteria (e.g., of people with hearing or vision problems, or language disorders), and likely undersampling of some groups (e.g., people without regular access to stable WiFi or personal electronic devices) may limit generalizability of findings. Since we aimed to capture data from participants in line with their personal work schedules, the time of day assessments were completed, and the time between assessments, was not controlled for. There may also have been individual shift work patterns (e.g., 24 hours on / 48 hours off) that were not well accommodated under our twice-daily assessment design. Time since sleep and time since work were calculated in hours based on users’ reporting of their most recent sleep and work end times (rather than e.g., automated logging of this data), so these measure are likely subject to some degree of measurement error. Finally, for the within-user analysis, we were required to filter the data for participants with a minimum number of unique time-point observations, which may have affected the profile of users included in this analysis (in particular, in a sample with slightly lower baseline depression scores compared to users who completed fewer time-points).

Overall, we argue that these findings offer preliminary support for the utility of using brief momentary speech samples as a low-burden, acceptable, reliable and repeatable measure of different aspects of fatigue relevant to high-risk populations. Future work should explore improvements in accuracy using more complex base models, whilst carefully considering the best way of taking into account differences in demographics across users, and the potential for capitalizing on user-specific models for less tractable but clinically important aspects of fatigue.

### Clinical implications

Fatigue has previously been estimated to be the cause 5-7% of primary care appointments in the UK [51]. The development of accurate and reliable digital measurement tools for fatigue is therefore likely to be of broad clinical relevance. Building understanding of complex multidimensional phenomena such as fatigue requires intensive longitudinal measurements within the same individuals: data that is hard to obtain using current methods [9]. Importantly, building such knowledge promises not only important research insights into how prolonged circadian disruption develops into risk for poorer long-term physical and mental health, but the possibility of returning actionable information directly to users. Returning individual fatigue results to users, supported by appropriate visualizations and contextualizing information (e.g., comparison to relevant peers and own past data), can act as prompts for reflection and action [52, 53], including the opportunity for just-in-time intervention [54]. For shift workers - including those in safety-critical industries such as construction, transportation and healthcare - early fatigue detection may help prevent accidents and other undesirable outcomes by supporting appropriate shift scheduling and timely recovery interventions such as rest breaks and naps. New fatigue measurement tools would help address an important gap in current provision, since previously existing measures (the UK’s Health and Safety Executive’s Fatigue Risk Index) have recently been withdrawn due to poor performance [55].

## Data availability

Due to licensing and IP considerations, the supporting dataset is not generally publicly available. However, we are open to partnering with research institutes and individual academics, including data sharing.

## Acknowledgments

We extend our gratitude to all the participants who generously dedicated their limited time and effort to our study.

## Funding

This work was supported by Innovate UK, the United Kingdom’s innovation agency, under Grant No. 10042845.

## Competing Interests

The authors declare that the research presented in this paper may be used in the development of products by the company. Thymia Limited has submitted applications for patents for digital health-tracking tools.

## Supplementary Methods

### Study flow

After reading the study description page on Prolific, and registering interest in the study, participants were routed to a thymia platform page where they could view the study information sheet and provided electronic consent. Upon consenting to take part in the study, participants were forwarded to their personal thymia dashboard, which lists currently scheduled study activities.

At the start of the study, participants completed a comprehensive demographic questionnaire, and brief measures of depression and anxiety symptoms, suitable for use in the general population (the PHQ-8 and GAD-7; [34, 35]).

During the main part of the study (two available assessments per day for 14 days), participants completed several brief speech elicitation activities, and provided information about their recent work and sleep activities and current fatigue levels (total time 1̃0 minutes). Activities were always presented in the same order on participants’ dashboards, but order of completion was not strictly enforced.

Participants were compensated for their time and effort at a rate of £9 per hour. Data were collected between November 2022 and February 2023.

### Speech activities

Three different speech activities (all 1̃ minute in length) were included at each study time-point: 1) “Read Aloud” (a paragraph reading activity), 2) “Read Aloud (Nonsense)” (a scrambled sentence reading activity), and 3) “Describe a Picture” (a spontaneous narration activity).

The “Read Aloud” task is commonly used in the scientific literature to elicit speech. Texts were taken from the previously-published speech literature and included the Aesop fables, The North Wind and the Sun, The Boy who Cried Wolf [56], and the Rainbow Passage [57]. The “Read Aloud (Nonsense)” task is an increased cognitive load reading task, which involves reversing the order of adjacent words in the previous texts to create scrambled sentences that are more difficult to read (see [58]). The “Describe a Picture” task is designed to elicit spontaneous narrative discourse [59]. In this version, participants were presented with rich and colourful illustrations, which included human and animal characters, actions happening in the foreground and background, and displays of mental states.

Participants were presented with 14 different variants of each activity (i.e., unique activities at each time-point for each week of the study), in order to minimise repetitiveness and maintain engagement. Specifically, participants saw different stimuli in the morning and evening of the the first week, until all 14 stimuli were seen once. In the second week of the study, stimuli were repeated, in reversed daily order to the previous week.

### Self-reported fatigue

Self-reported fatigue was measured using 10-point likert scales Sleepiness was measured using the item “*Please select the point on the scale below which best describes your sleepiness level during the last 10 minutes from 1 = extremely alert and awake to 10 = extremely sleepy, can’t keep awake*”. Physical exhaustion was measured using the item “*I feel physically exhausted or weary from putting effort into an activity (like a chore or a shift)*”. Mental exhaustion was measured using the item “*I feel mentally exhausted or weary from putting effort into an activity (like a chore or a shift)*”. Lack of energy or anergia was measured using the item “*I feel sluggish, drained of energy or lacking strength without having exerted myself*”. Lack of motivation was measured using the item “*I feel unmotivated to do anything currently - but this is NOT due to feeling tired, exhausted. I just feel like I can’t be bothered*”.

## Supplementary Results

### Differences in variances and intercepts in fatigue measures over the course of the study

Within-user standard deviations over the course of the study were significantly lower for self-reported fatigue measures than time since sleep (*t*_1075_=11.7, 14.9, 13.3, 16.0, 20.5 for sleepiness, physical exhaustion, mental exhaustion, lack of energy, and lack of motivation, respectively; all *p <* 0.001 paired-samples *t*-tests based on within-user SDs of normalized fatigue scores, in all training data). This indicates that within-user variances in subjective fatigue ratings were lower than within-user variances in time since sleep over the study period.

Between-user variances in mean fatigue measure values (averaged over all time-points) were also significantly higher for self-reported fatigue scores, compared to time since sleep (*F*_1075_=1.70, 2.06, 1.90, 2.21, 2.35 for sleepiness, physical exhaustion, mental exhaustion, lack of energy, and lack of motivation, respectively; all *p <* 0.001 *F* tests for equal variances over within-user means of normalized fatigue scores, in training data). This indicates that within-user mean (intercept) scores were more variable for subjective fatigue ratings than for time since sleep measures over the course of the study.

### Differences in between users included in cross-sectional and within-user analysis

For the within-user analysis, the training dataset was filtered for users with data from at least six different study-timepoints. Since continued participation in the study may vary according to individual circumstances, we investigated if users in the filtered dataset differed from the users who provided fewer time-point observations in terms of baseline sociodemographic and clinical measures.

Users who completed 6 study time-points or more were slightly older (*t*_945_ = 2.83*, p* = 0.005, mean age 34.3 vs 32.6) and were more likely to be Spanish language speakers (*χ*^2^ = 34.2*, p <* 0.001, proportions = 0.24 vs 0.10), but did not differ in terms of birth sex (*χ*^2^ = 5.41*, p* = 0.07). Users with 6 or more time-points had lower baseline PHQ-8 total scores (*t*_901_ = 3.64, mean total score 7.8 vs 9.1), potentially reflecting lower ability to engage with repeated measurement prompts in more depressed users.

---

**Table S1:**
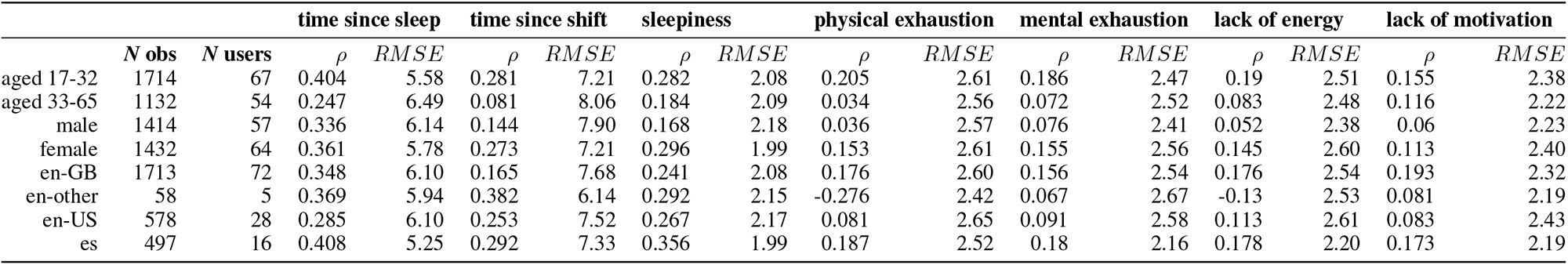
Cross-sectional model performance in the independent test set by demographic group (single speech activity models). Performance is reported separately by age, birth sex, and language/accent categories. *N obs*, total number of observations for that category in test data. *N users*, total number of unique users for that category in test data.

**Table S2:**
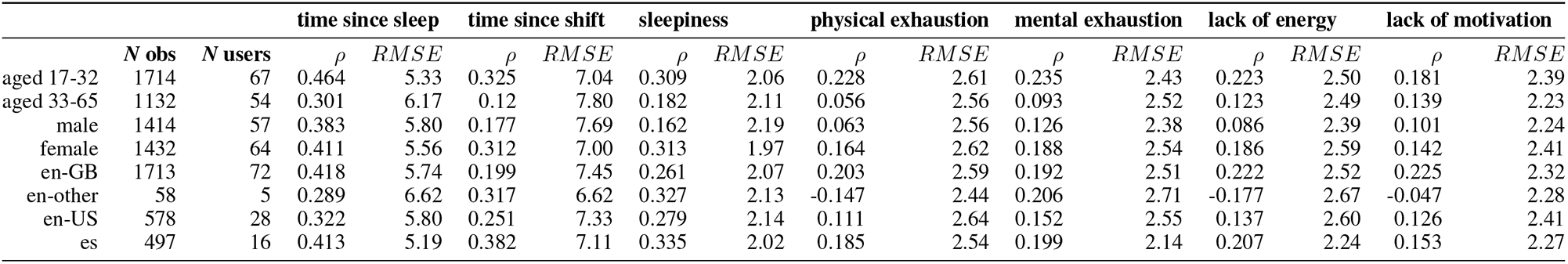
Cross-sectional model performance in the independent test set by demographic group (averaged of speech activity models). Performance is reported separately by age, birth sex, and language/accent categories. *N obs*, total number of observations for that category in test data. *N users*, total number of unique users for that category in test data.

**Table S3:**
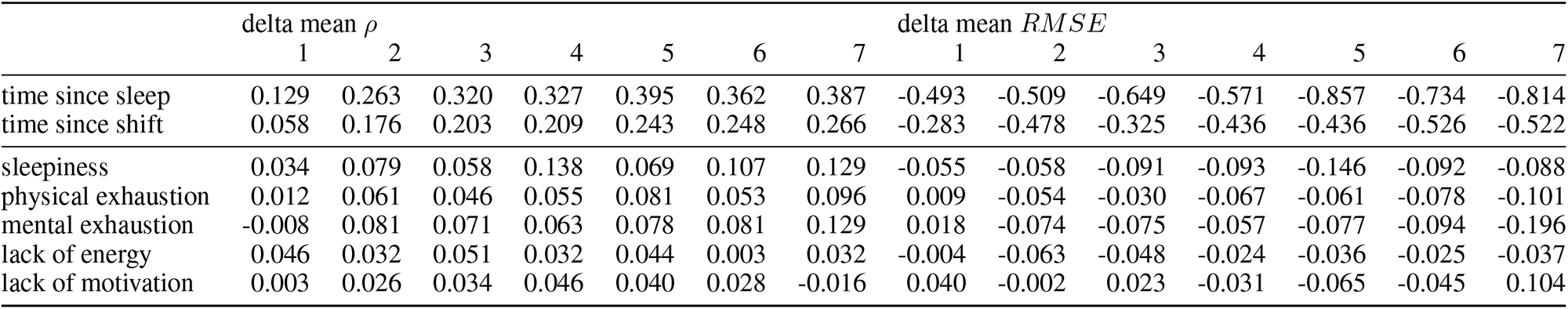
Within-user model performance, by number of included training observations from the same user (single activity model). Delta values represent differences in performance between the model including speech features and the relevant null (intercept only) model: with increases in *ρ* (positive values) and decreases *RMSE* (negative values) representing superior performance for the speech model.

**Table S4:** Within-user model performance, by number of included training observations from the same user (averaged over activities model). Delta values represent differences in performance between the model including speech features and the relevant null (intercept only) model: with increases in *ρ* (positive values) and decreases *RMSE* (negative values) representing superior performance for the speech model.

| | delta mean $\rho$ | | | | | | | delta mean $RMSE$ | | | | | | |
| --- | --- | --- | --- | --- | --- | --- | --- | --- | --- | --- | --- | --- | --- | --- |
|  | 1 | 2 | 3 | 4 | 5 | 6 | 7 | 1 | 2 | 3 | 4 | 5 | 6 | 7 |
| time since sleep | 0.114 | 0.269 | 0.325 | 0.328 | 0.355 | 0.357 | 0.404 | -0.653 | -0.701 | -0.712 | -0.665 | -0.761 | -0.829 | -0.892 |
| time since shift | 0.078 | 0.166 | 0.210 | 0.199 | 0.206 | 0.255 | 0.282 | -0.295 | -0.357 | -0.427 | -0.404 | -0.320 | -0.478 | -0.449 |
| sleepiness | 0.025 | 0.062 | 0.052 | 0.139 | 0.047 | 0.080 | 0.149 | -0.224 | -0.151 | -0.094 | -0.162 | -0.144 | -0.068 | -0.125 |
| physical exhaustion | 0.028 | 0.041 | 0.049 | 0.035 | 0.087 | 0.051 | 0.098 | -0.256 | -0.147 | -0.132 | -0.094 | -0.071 | -0.084 | -0.134 |
| mental exhaustion | -0.004 | 0.048 | 0.030 | 0.039 | 0.067 | 0.107 | 0.105 | -0.174 | -0.180 | -0.077 | -0.030 | -0.061 | -0.093 | -0.122 |
| lack of energy | 0.026 | 0.013 | 0.050 | 0.019 | 0.035 | 0.015 | 0.034 | -0.216 | -0.114 | -0.087 | -0.066 | -0.102 | -0.027 | -0.028 |
| lack of motivation | 0.006 | -0.009 | 0.004 | 0.060 | 0.009 | 0.054 | 0.001 | -0.131 | -0.088 | -0.036 | -0.037 | -0.067 | -0.087 | 0.061 |

**Figure S1:**
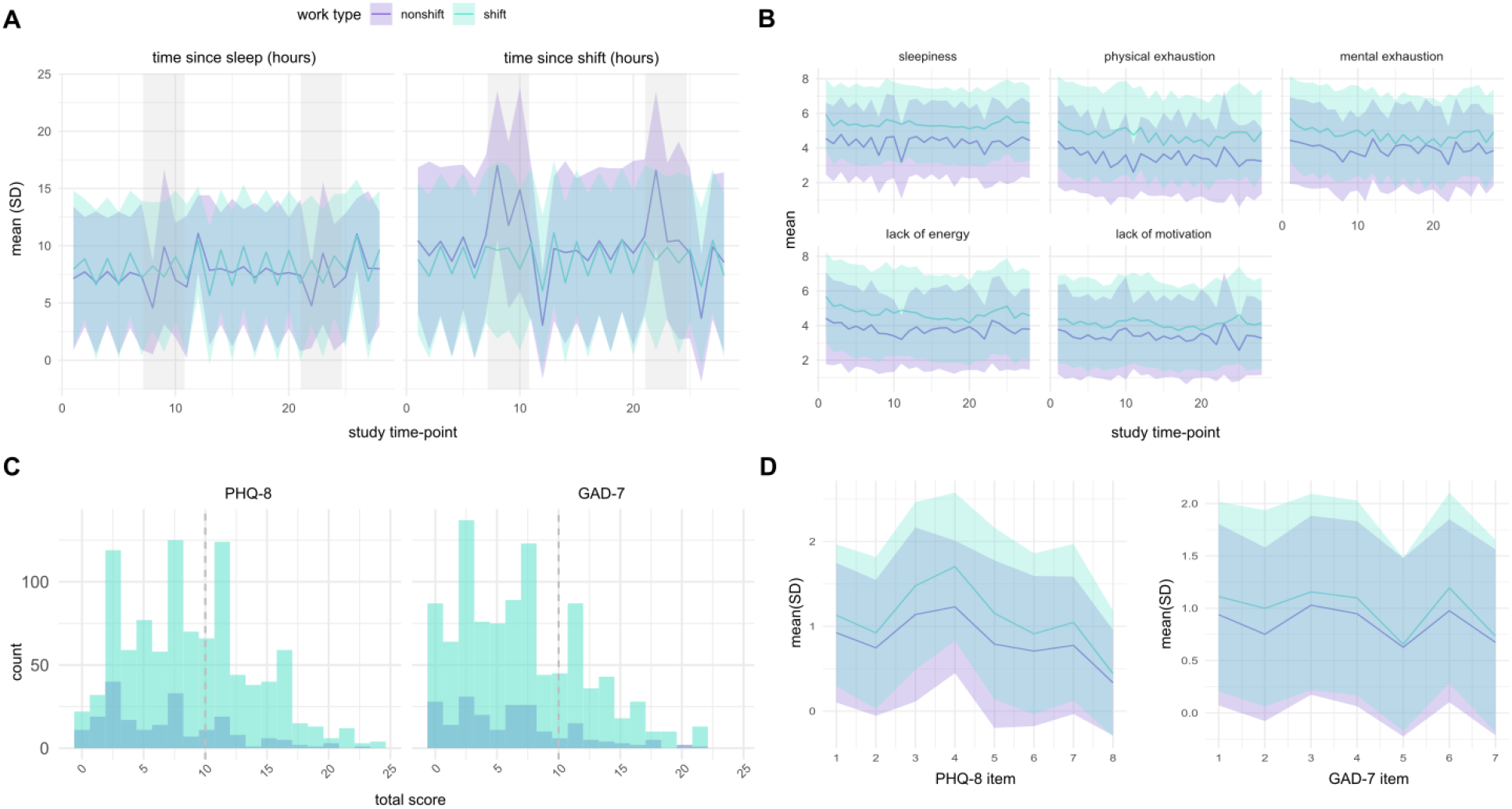
Differences in objective and subjective fatigue, depression, and anxiety symptoms by shift work status. **A** Mean (SD) time since sleep and and time since shift over the course of the study, by shift work status. Grey shading represents weekends. **B** Mean (SD) subjective fatigue ratings over the course of the study, by shift work status. **C** Distributions of baseline total PHQ-8 depression scores and GAD-7 anxiety scores, by shift work status. Grey dotted lines represent accepted cut-offs for clinically-significant levels of depression and anxiety symptoms. **D** Differences in mean baseline scores between shift and non-shift workers in individual PHQ-8 and GAD-7 scores.

**Figure S2:**
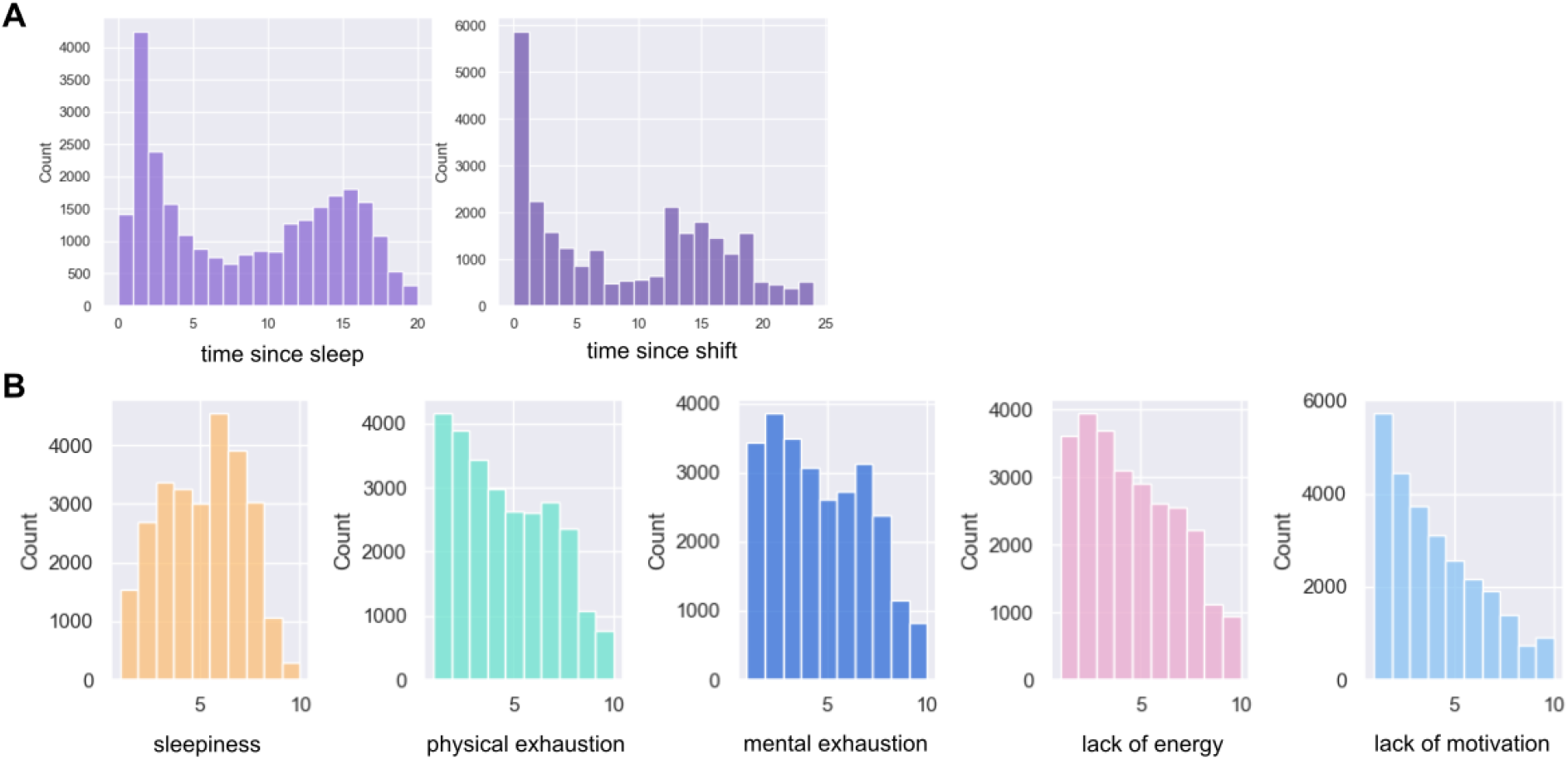
Distribution of observed fatigue measures in the training data set. **A** Time since sleep and time since shift (in hours). **B** Subjective fatigue ratings (possible scores 0-10).

**Figure S3:**
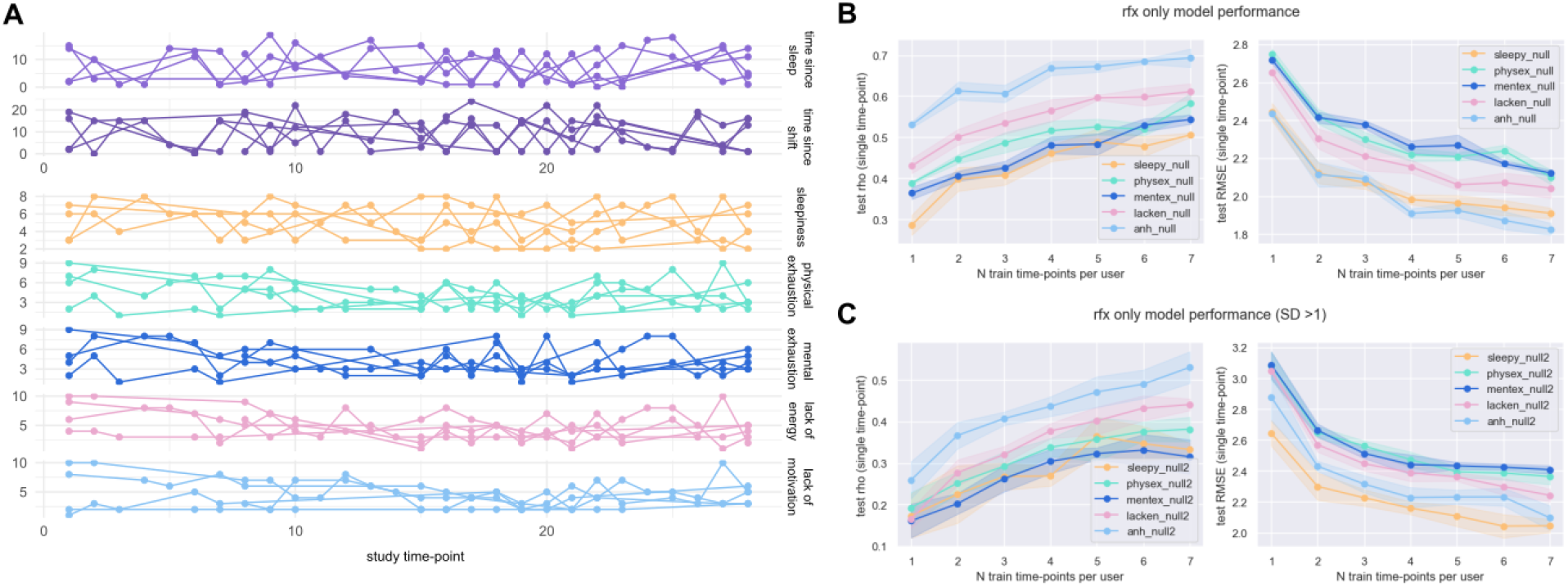
Differences in within-user objective and subjective fatigue data. **A** Time since sleep, time since shift, and self-reported subjective fatigue levels over the course of the study for 5 randomly-selected users. Of note, the subjective fatigue measures move more slowly over the course of the study and show stronger user-specific intercept effects. **B** Null (intercept-only) model performance for subjective fatigue ratings across all users with >=6 different observed time-points. **C** Null (intercept-only) model performance for subjective fatigue ratings across all users with >=6 different observed time-points, filtered for >=1SD in variance over study time-points.

**Figure S4:**
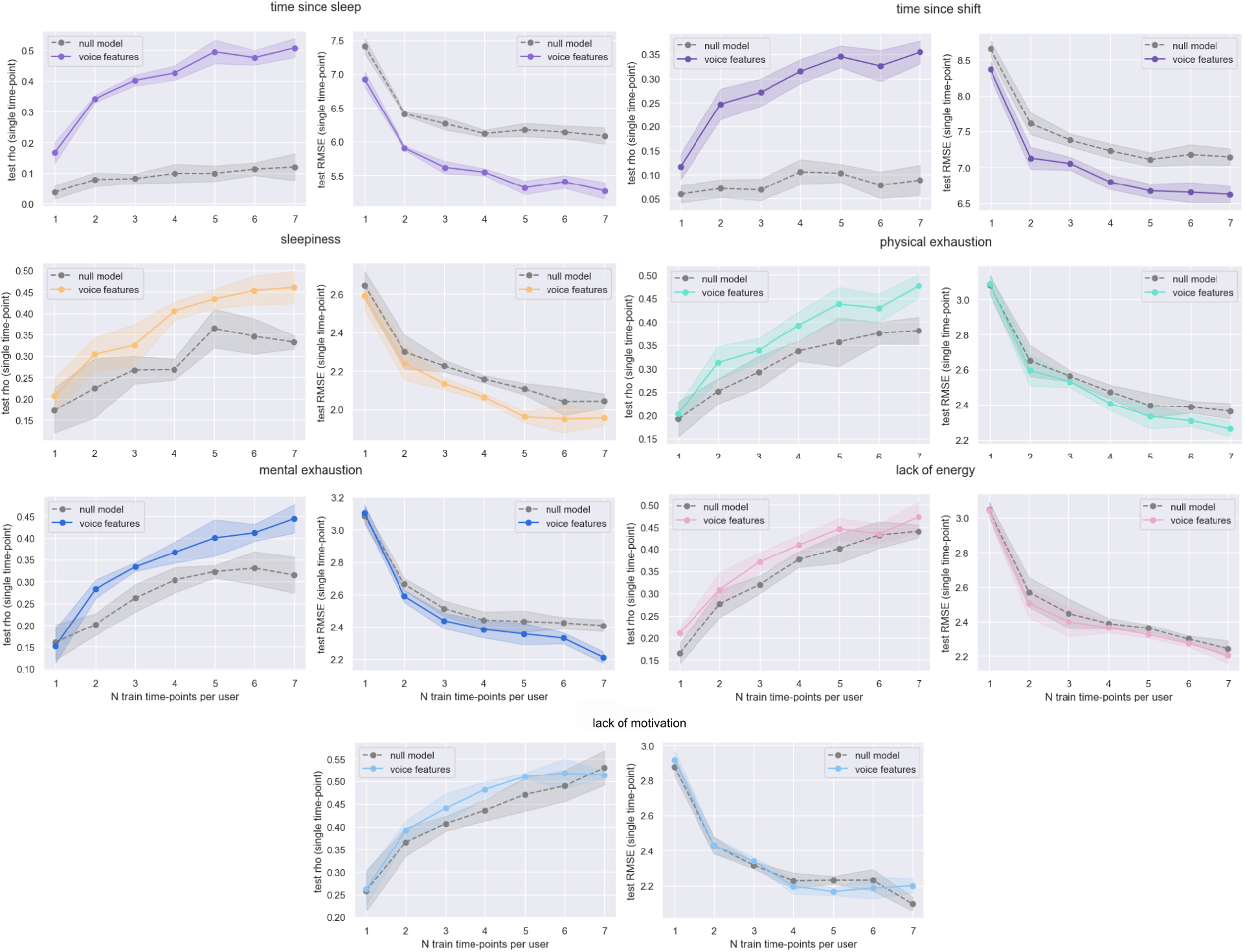
Within-user model performance, for the single activity model. For each fatigue measure, performance for a single unseen time-point is plotted against number of observations from the same user included in the training data. Speech feature models included all speech features at each time point, cluster (user) ID, and random-effects. Null models included only cluster (user) ID and random-effects values. Rho values are Spearman correlation coefficents between predicted and observed values in independent test set data. RMSE, root mean squared error of predictions.

## Notes

### Author Declarations

Ethical approval for study procedures was supplied by Dr David Carpenter, an external research ethics consultant identified through the Association of Research Managers and Administrators (ARMA, https://arma.ac.uk/) as part of a commercial ethical review process. Ethical approval was granted on 22nd November 2023.

